# Instability of high polygenic risk classification and mitigation by integrative scoring

**DOI:** 10.1101/2024.07.24.24310897

**Authors:** Anika Misra, Buu Truong, Sarah M. Urbut, Yang Sui, Akl C. Fahed, Jordan W. Smoller, Aniruddh P. Patel, Pradeep Natarajan

## Abstract

Polygenic risk scores (PRS) continue to improve with novel methods and expanding genome-wide association studies. Healthcare and commercial laboratories are increasingly deploying PRS reports to patients, but it is unknown how the classification of high polygenic risk changes across individual PRS. Here, we assessed association and classification performance of cataloged PRS for three complex traits. We chronologically ordered all trait-related publications (Pub_n_) and identified the single PRS Best(Pub_n_) for each Pub_n_ that had the strongest association with the target outcome. While each Best(Pub_n_) demonstrated generally consistent population-level strengths of associations, classification of individuals in the top 10% of each Best(Pub_n_) distribution varied widely. Using the PRSmix framework, which integrates information across several PRS to improve prediction, we generate corresponding ChronoAdd(Pub_n_) scores for each Pub_n_ that combine all polygenic scores from all publications up to and including Pub_n_. When compared with Best(Pub_n_), ChronoAdd(Pub_n_) scores demonstrated more consistent high-risk classification amongst themselves. This integrative scoring approach provides stable and reliable classification of high-risk individuals, and is an adaptable framework into which new scores can be incorporated as they are introduced, integrating easily with current PRS implementation strategies.

## Main

Polygenic risk scores (PRS), which quantify the genetic risk for traits from common variants, have improved in their predictive performances over the past decade.^1^ Building on classical approaches of pruning and thresholding, methods incorporating Bayesian approaches using prior knowledge about genetic architecture, relatedness of individuals, linkage disequilibrium patterns, and genetic effects across populations have improved assignment of variant weights and PRS performance.^2–5^ In parallel, genome-wide association studies (GWAS) have continued to expand in size, with the most recent iteration of the GWASs for human height and coronary artery disease reaching 5.4 million and 1.3 million participants, respectively.^6,7^ Incorporation of multi-ancestry and multi-trait GWAS data has further improved PRS prediction in diverse ancestral groups.^8^ In attempting to quantify total inherited risk, each of these PRS iterations for a given trait captures unique information contingent on source GWAS, method, and training dataset.

PRS are increasingly being delivered to patients. Third-party genetic testing companies and healthcare system laboratories are already delivering polygenic scores for coronary artery disease, diabetes, cancers, and other diseases.^9,10^ The eMERGE consortium has developed PRS reports for 10 diseases to return to participants within healthcare system as part of a larger effort to study genomic risk assessment and management.^11–13^ Furthermore, researchers recently developed clinically valid assays, clinical workflows, and patient– and physician-oriented information materials to accompany PRS reports delivered within the Mass General Brigham Biobank and the Veterans Affairs Genomic Medicine at Veterans Affairs (GenoVA) Study.^14,15^ New clinical trials are incorporating PRS into medical decision-making (NCT05819814, NCT05850091), and medical societies have begun to release initial statements on their utility.^16,17^

Although consistency in risk classification is important for clinical decision making, risk prediction models for complex disease will inherently have variability in high-risk classification. These differences often stem from differences in training population, outcome definition, and underlying statistical methodology. Introduction of new models often leads to significant reclassification of high risk among patients, raising questions of need for altering recommendations for interventions or therapies. For example, since 2013, the American College of Cardiology and the American Heart Association (AHA) have recommended using the Pooled Cohort Equations (PCE)^18^ to estimate 10-year risk for atherosclerotic cardiovascular disease, but the AHA recently developed the Predicting Risk of CVD EVENTs (PREVENT) equations,^19^ which include kidney measures, exclude race, and include heart failure in the composite predicted outcome. When applied to representative cohorts, PREVENT predicts lower cardiovascular risk than PCE and could reclassify about half of US adults to lower risk categories, potentially leading to reduced statin and antihypertensive treatment eligibility and increasing incidence of composite cardiovascular outcomes.^20^ Similarly, current reports of polygenic risk also provide a categorical assessment of high risk, and different studies and companies use different scores for the same traits.^9–11^ As advances in GWAS size and statistical methods continue to fuel release of improved new PRS, a framework is needed on updating scores or incorporating new data without causing confusion.

Despite ongoing progress toward clinical implementation, the variability of individual-level classification of ‘high genetic risk’ using different PRS for a given trait remains largely untested, and consensus PRS for any trait currently do not exist. Previous efforts have focused on population-level prediction metrics, rather than consistency of high-risk classification presented in individual clinical reports now as a part of clinical implementation workflows.^8^ Prior limited availability of large holdout diverse datasets has precluded individual-level benchmarking to assess agreement in classification between PRS. Furthermore, there is a need to aggregate and incorporate orthogonal data from available PRS while overcoming correlation between scores and maximize predictive performance.

Using the large and ancestrally-diverse *All of Us* (AOU) cohort^21^, we set out to compare the classification of individuals with high genetic risk based on published polygenic scores for three common, complex diseases: coronary artery disease (CAD), type 2 diabetes (T2DM), and major depressive disorder (MDD). We also test the effect of using PRSmix^22^ — a tool that agnostically integrates information across several PRS for a given trait to improve prediction accuracy for a target population — in influencing high genetic risk classification over iterations of polygenic scores.

## Results

We determined the associations of published PRS for complex traits from the Polygenic Score Catalog^23^ in AOU, which has aggregated genotype data and extensive phenotypic information on 236,393 participants (average enrollment age: 51.8 years; 60.6% female; genetically inferred ancestry of 54.7% EUR, 22.9% AFR, 18.9% AMR, 2.3% EAS and 1.1% SAS).^24^ Specifically, we calculated 57 scores for CAD, 129 scores for T2DM, and 18 scores for MDD.(Supplemental Tables 1-3) We tested the associations of these scores with corresponding outcomes. Some publications deposited multiple trait-specific scores in the PGS Catalog: the 57 CAD scores were published by a total of 40 unique publications. Similarly, the 129 T2DM scores were deposited by only 39 unique publications, and the 18 MDD scores were deposited by 7 publications. Because we sought to evaluate the comparative differences of the successively newly available PRS, we ordered the publications of each trait chronologically based on the source publication date, enumerating them as Pub_n_. In a given trait, for each publication Pub_n_ in the ordered list of all trait-specific Pub_n_, we then identified the dominating PRS – i.e. the PRS that associated most strongly with the target disease – deposited by the given Pub_n_ to carry forward in subsequent analysis, labeling it the Best(Pub_n_) with *n* referring to the chronological order of Pub_n_ in the list of all available trait-specific publications. This resulted in 40 CAD Best(Pub_n_), 39 T2DM Best(Pub_n_), and 7 MDD Best(Pub_n_).(Figure 1)

**Figure 1:**
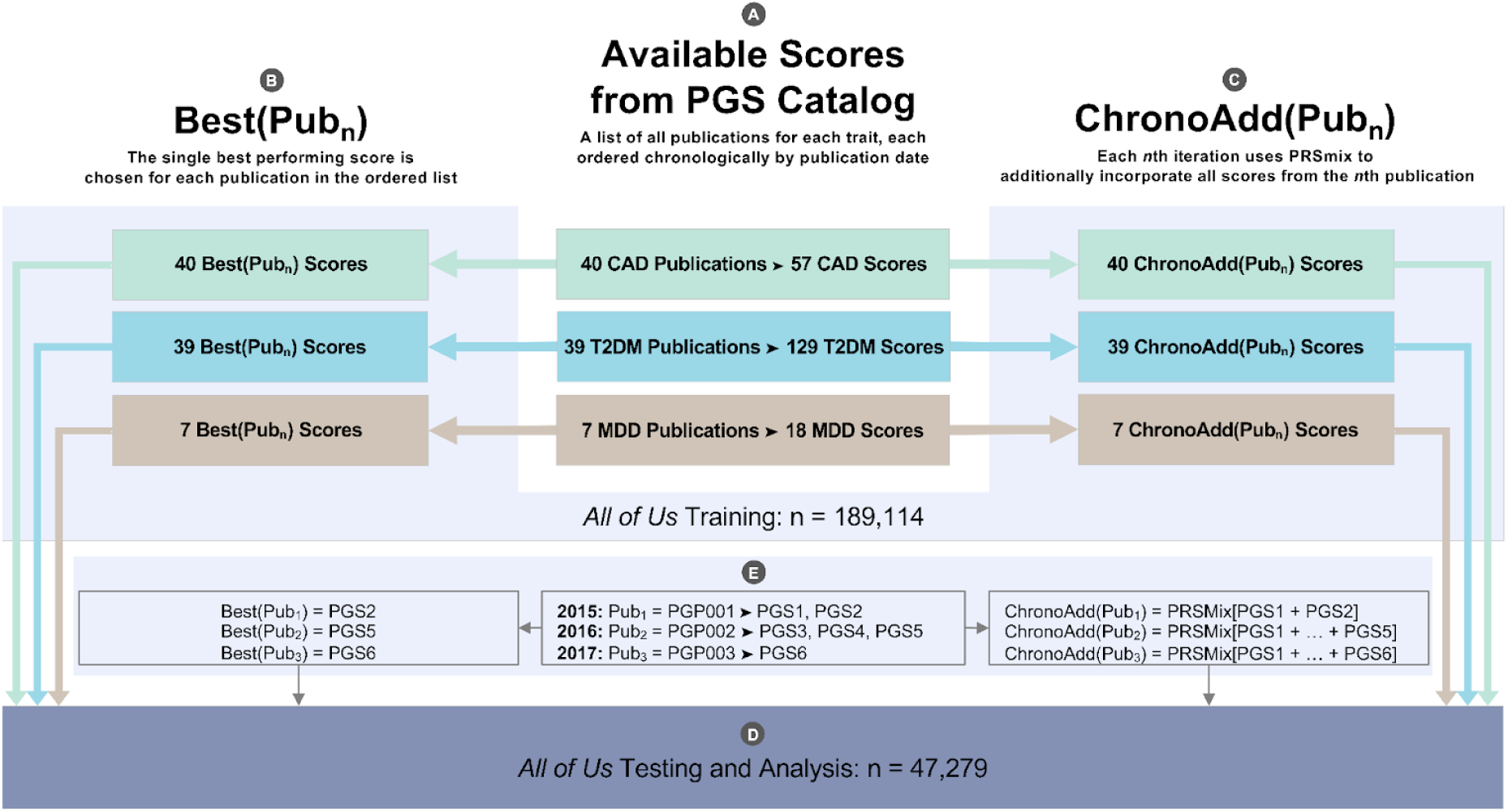
Overview of analysis. A) All polygenic scores that do not include *All of Us* (AOU) participants in their development were downloaded from the PGS catalog and calculated in AOU for three traits of interest: coronary artery disease (CAD), type 2 diabetes mellitus (T2DM), and major depressive disorder (MDD). Many publications deposited several scores, resulting in 40 publications depositing a total of 57 single scores for CAD, 39 publications depositing 129 scores for T2DM, and 7 publications depositing 18 scores for MDD. For each trait, all the score-depositing publications were ordered chronologically by publication date. B) Within each publication Pub_n_ for a given trait, the strongest score deposited by the publication – henceforth referred to as Best(Pub_n_) – was chosen as the score representative of the publication. This yielded a chronologically ordered list of Best(Pub_n_) corresponding to the list of Pub_n_ for each trait. C) For each publication Pub_n_ in the chronologically ordered list of publications of a given trait, PRSmix was applied in the training set to generate a corresponding ChronoAdd(Pub_n_) score by mixing all the scores from Pub_n_ with all the other scores from all the previous publications in the list: Pub_1_ through Pub_n-1_. D) A 1:1 comparative analysis was performed in chronological order between the pairs of corresponding Best(Pub_n_) and ChronoAdd(Pub_n_) for each publication Pub_n_ of each trait in the holdout cohort of 47,279 AOU individuals. E) For example, consider a trait with three associated PGS publications depositing multiple scores. Per (A), the publications are ordered by publication date, resulting in PGP001, PGP002, and PGP003 being labeled as Pub_1_, Pub_2_, and Pub_3_, respectively. Per (B), Best(Pub_n_) are the single best scores for each of these publications based on performance in the training cohort, resulting in PGS2, PGS5, and PGS6 being labeled as Best(Pub_1_), Best(Pub_2_), and Best(Pub_3_), corresponding to Pub_1_, Pub_2_, and Pub_3_, respectively. Per (C), ChronoAdd(Pub_n_) is calculated by mixing all scores from all publications up to and including Pub_n_: ChronoAdd(Pub_1_) = PRSmix[Pub_1_] = PRSmix[PGS1 + PGS2], ChronoAdd(Pub_2_) = PRSmix[Pub_1_ + Pub_2_] = PRSmix[PGS1 + PGS2 + PGS3 + PGS4 + PGS5], and so on.

### Assessment of high-risk classification stability between polygenic scores

First, we calculated the associations and classification stability of high polygenic risk, defined as the top 10% of the score distribution, with the corresponding trait. Using contemporary PRS for common diseases, defining a threshold of top 10% as high genetic risk is associated with approximately two-fold greater risk of disease in prior studies.^14^ The ORs associated with top 10% risk classification ranged from 1.25-2.50 for all CAD Best(Pub_n_) with CAD, 1.30-2.58 for all T2DM Best(Pub_n_) with T2DM, and 0.99-1.47 for all MDD Best(Pub_n_) with MDD.(Supplemental Tables 4-6) While each Best(Pub_n_) demonstrated consistent strengths of association, classification of individuals into the top 10% of each PRS distribution varied widely. The Jaccard index, a metric of similarity, is the proportion of observations in agreement between two sets of data relative to the total number of observations. The median [interquartile range] Jaccard index for top 10% classification by CAD Best(Pub_n_) was 0.17 [0.13-0.22], indicating poor agreement across scores. Similarly, the median Jaccard index for the top 10% classification was 0.18 [0.15-0.22] for T2DM Best(Pub_n_) and 0.11 [0.08-0.17] for MDD Best(Pub_n_) (Figure 2). This trend was invariant of threshold choice. (Supplemental Table 7) When restricting to polygenic scores from the five most recent publications, we continued to observe poor agreement of high-risk classifications, with median [IQR] Jaccard index estimates of 0.15 [0.13-0.22] for CAD Best(Pub_n_), 0.26 [0.18-0.35] for T2DM Best(Pub_n_), and 0.18 [0.07-0.23] for MDD Best(Pub_n_).(Supplemental Table 7)

**Figure 2:**
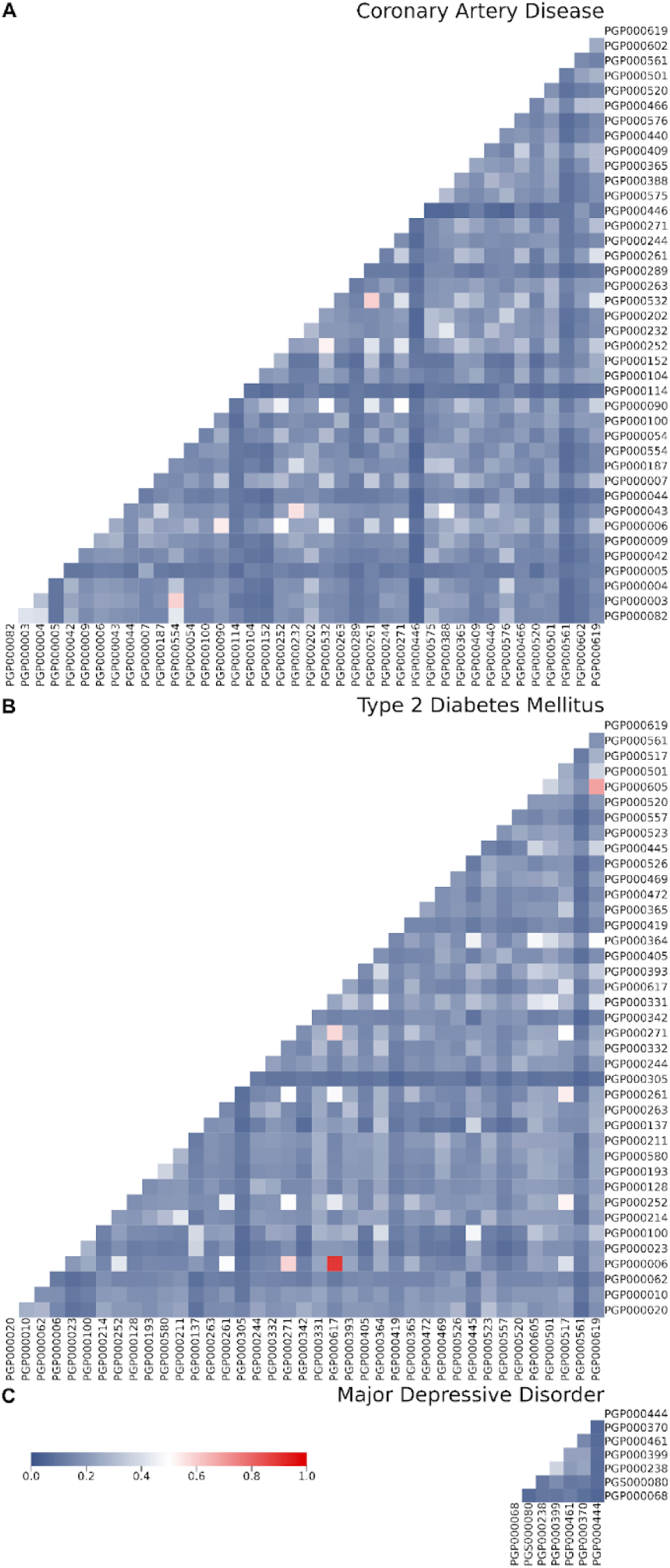
Jaccard index heatmaps for high genetic risk across trait-specific Best(Pub_n_) pairs. Jaccard indices of similarity calculated for pairs of classification of top 10% risk determined by Best(Pub_n_) polygenic scores for coronary artery disease, type 2 diabetes mellitus, and major depressive disorder. Scores are ordered chronologically based on date of publication, advancing left to right and bottom to top. The median [interquartile range] Jaccard index for top 10% classification was: A) 0.17 [0.13-0.22] for coronary artery disease; B) 0.18 [0.15-0.22] for type 2 diabetes mellitus; and C) 0.11 [0.08-0.17] for major depressive disorder.

### Risk classification in simulation studies of correlated distributions

The behavior of correlated variables at the extremes of distributions can differ from their behavior in the middle. In most biological distributions, such as the multivariate normal distribution, the correlation at the extremes tends to be lower, a phenomenon known as tail independence in extreme value theory.^25^ We tested this principle by generating simulated correlated distributions and assessing the correlations of their extremes. By modulating the specified strength of the overall correlation of the simulated distributions at expected Pearson correlation indices of *r*=0.2, 0.5, and 0.8, we noted correlations in the middle of the distributions to be 0.202, 0.502, and 0.802, and correlations in the high extremes of these distributions to be 0.087, 0.231, and 0.486.(Figure 3) This trend of significantly decreased correlation of high extremes was consistent across the full spectrum of overall correlation inputs.(See Supplemental Table 8) As greater overall correlation in scores was linked to higher correlation in the extremes, we hypothesized that an integrative scoring approach that incorporates elements of the most informative polygenic scores would lead to stronger associations, higher correlation, and greater consistency in high-risk classification across score versions.

**Figure 3:**
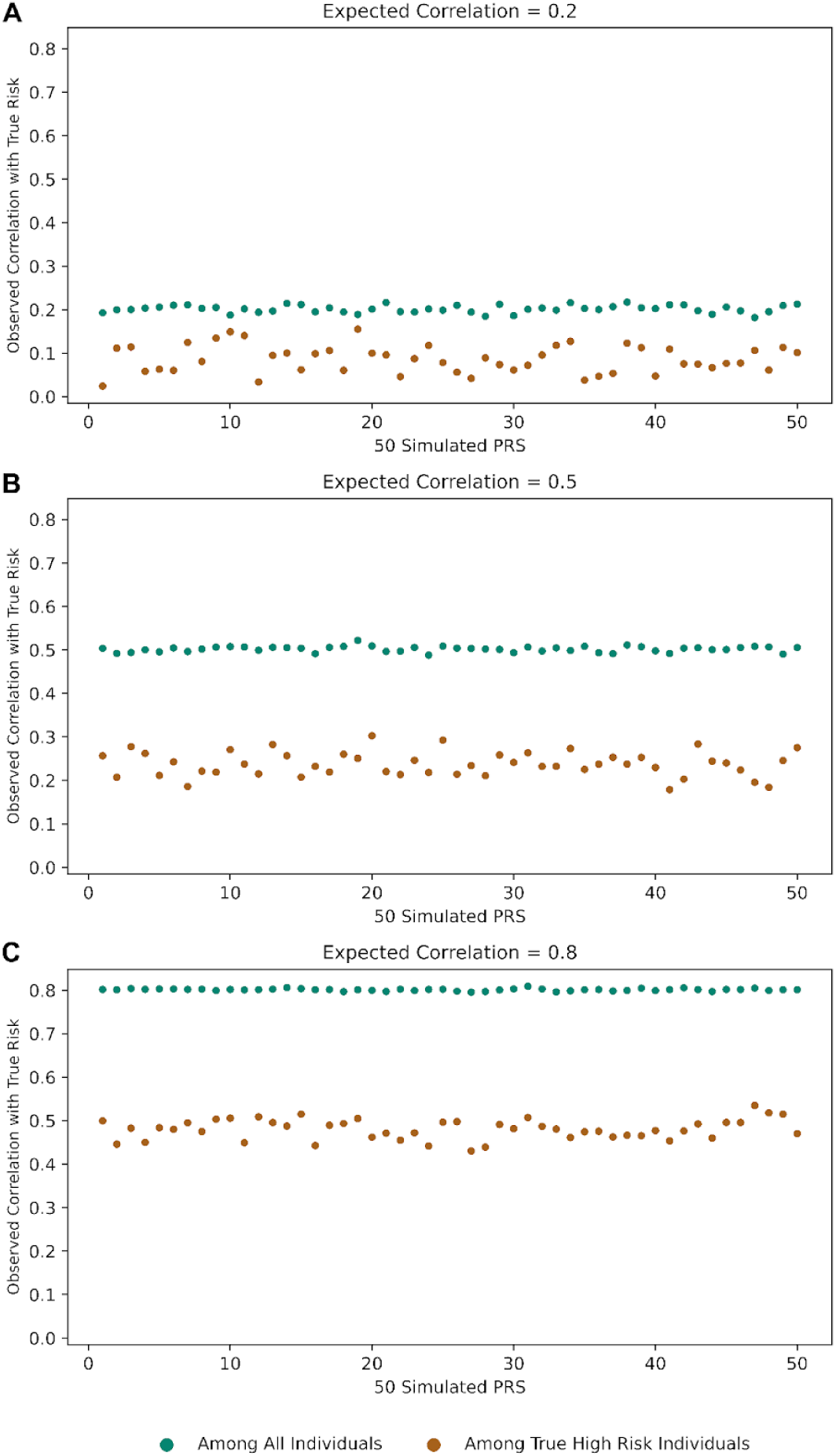
Simulations of correlated distributions in total population compared to in high extremes. A ground truth risk score for 10,000 individuals was simulated. For three different Pearson’s correlation coefficient values, 50 scores were simulated, where each score was expected to have a correlation with the true risk of approximately *r*. For each of these 50 scores, a correlation of the score with the true risk was calculated among all individuals and among only the high risk individuals (defined by those with a true risk in the top 10%). The observed correlation calculated only in high risk individuals is substantially lower, especially as correlation increases. A) For an expected correlation among all individuals of 0.2, the mean correlation with true risk across all 50 scores is 0.20 compared to 0.09 in high risk individuals only. B) Similarly, the mean correlation is 0.50 vs. 0.23 with an expected *r* of 0.5, and C) 0.80 vs. 0.49 with an expected *r* of 0.8.

### Integrative approach to develop ChronoAdd(Pub_n_) polygenic scores

We next used an integrative scoring approach to create sequential iterations of integrated polygenic scores. PRSmix is a software that takes in any number of trait-specific scores and combines them using an elastic net approach, outputting a new score that takes factors such as inter-score correlation and association strength into account to improve overall prediction accuracy.^22,26^ For a given trait, we generated a corresponding integrated score ChronoAdd(Pub_n_) for each publication Pub_n_, wherein all the scores deposited by the Pub_n_ in question were combined with all the scores from all the publications preceding it using PRSmix. We adopted a chronological approach in ordering based on publication date in order to retroactively demonstrate how the proposed integrative scoring approach would have worked in the past. With each new trait-relevant publication, we generated a new ChronoAdd(Pub_n_) score incorporating the new scores, mimicking the iterative chronological PRS revision, and the prospective process of healthcare systems incorporating new polygenic scores into their reporting framework as they are published. This allows for direct, temporal comparison between the best PRS in a given publication, Best(Pub_n_), with an integrated PRS that incorporates all trait-specific scores up to and including that publication, ChronoAdd(Pub_n_). We calculated a ChronoAdd(Pub_n_) for each publication in each trait, resulting in 40 CAD ChronoAdd(Pub_n_), 39 T2DM ChronoAdd(Pub_n_), and 7 MDD ChronoAdd(Pub_n_). For each trait, we selected the most recent ChronoAdd(Pub_n_), referred to as ChronoAdd(Pub_Last_), as the target for benchmarking the predictive performance of and high-risk classification by prior Best(Pub_n_) and ChronoAdd(Pub_n_) scores. Individuals in the top 10% of the ChronoAdd(Pub_Last_) distribution were defined as having true high genetic risk.

### Benchmarking high-risk classification using ChronoAdd(Pub_n_) polygenic scores

Using ChronoAdd(Pub_n_) resulted in more congruent high-risk classification compared to the respective Best(Pub_n_) itself over time. Using ChronoAdd(Pub_n_) the Jaccard index improved to median [IQR] of 0.39 [0.25-0.51] for CAD ChronoAdd(Pub_n_), 0.64 [0.45-0.83] for T2DM ChronoAdd(Pub_n_), and 0.46 [0.16-0.67] for MDD ChronoAdd(Pub_n_). (Figure 4) When restricting to ChronoAdd(Pub_n_) from the five most recent publications, we observed significantly higher congruence in high-risk classifications between ChronoAdd(Pub_n_) scores, with median [IQR] Jaccard index estimates of 0.76 [0.75-0.92] for CAD ChronoAdd(Pub_n_), 0.74 [0.74-0.80] for T2DM ChronoAdd(Pub_n_), and 0.67 [0.61-0.93] for MDD ChronoAdd(Pub_n_). (Figure 5) These findings of improved congruence for high genetic risk of CAD, T2DM, and MDD generalize across ancestral subgroups.(Figure 6)

**Figure 4:**
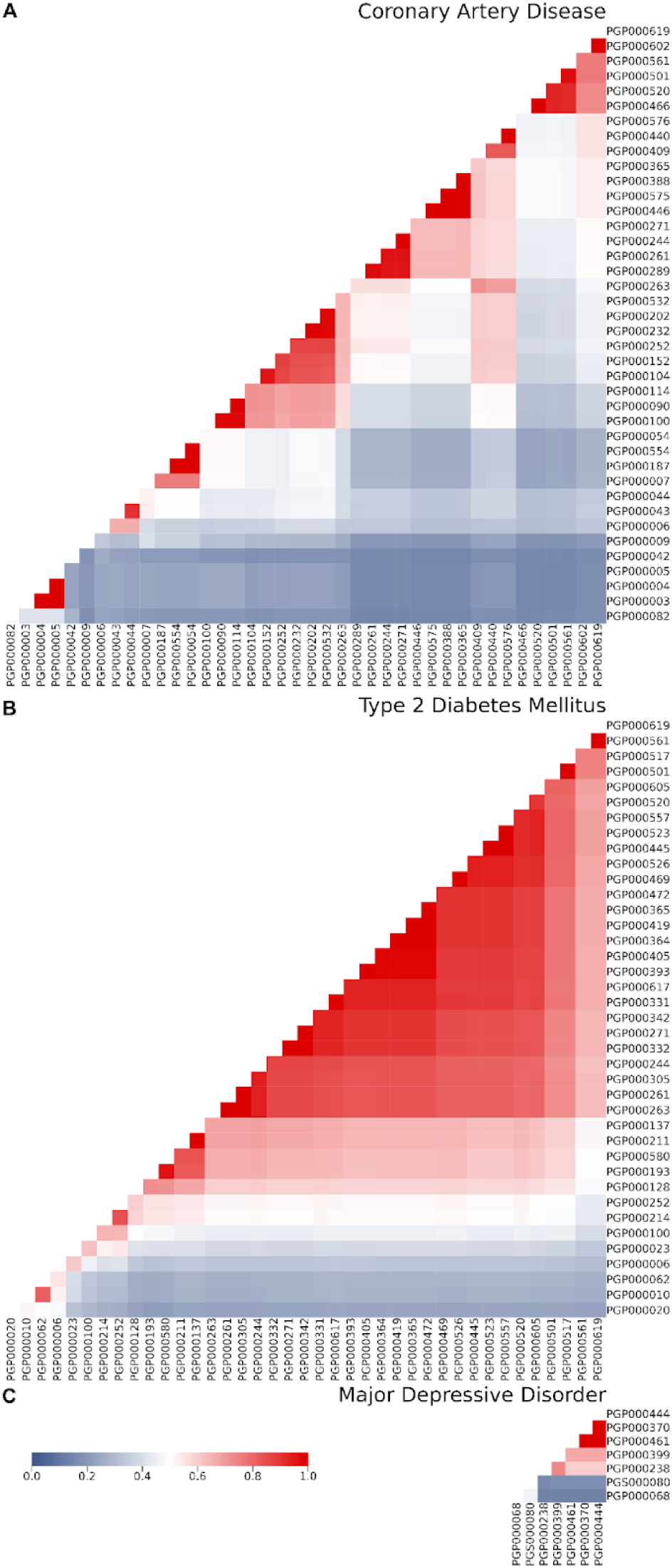
Jaccard index heatmaps for high genetic risk across trait-specific ChronoAdd(Pub_n_) pairs. Jaccard indices of similarity calculated for pairs of classification of top 10% risk determined by ChronoAdd(Pub_n_) polygenic scores generated by chronologically adding in all scores for each publication Pub_n_ using PRSmix framework for coronary artery disease, type 2 diabetes mellitus, and major depressive disorder. Scores are ordered chronologically based on date of publication, advancing left to right and bottom to top. The median [interquartile range] Jaccard index for top 10% classification was: A) 0.39 [0.25-0.51] for coronary artery disease; B) 0.64 [0.45-0.83] for type 2 diabetes mellitus; and C) 0.46 [0.16-0.67] for major depressive disorder.

**Figure 5:**
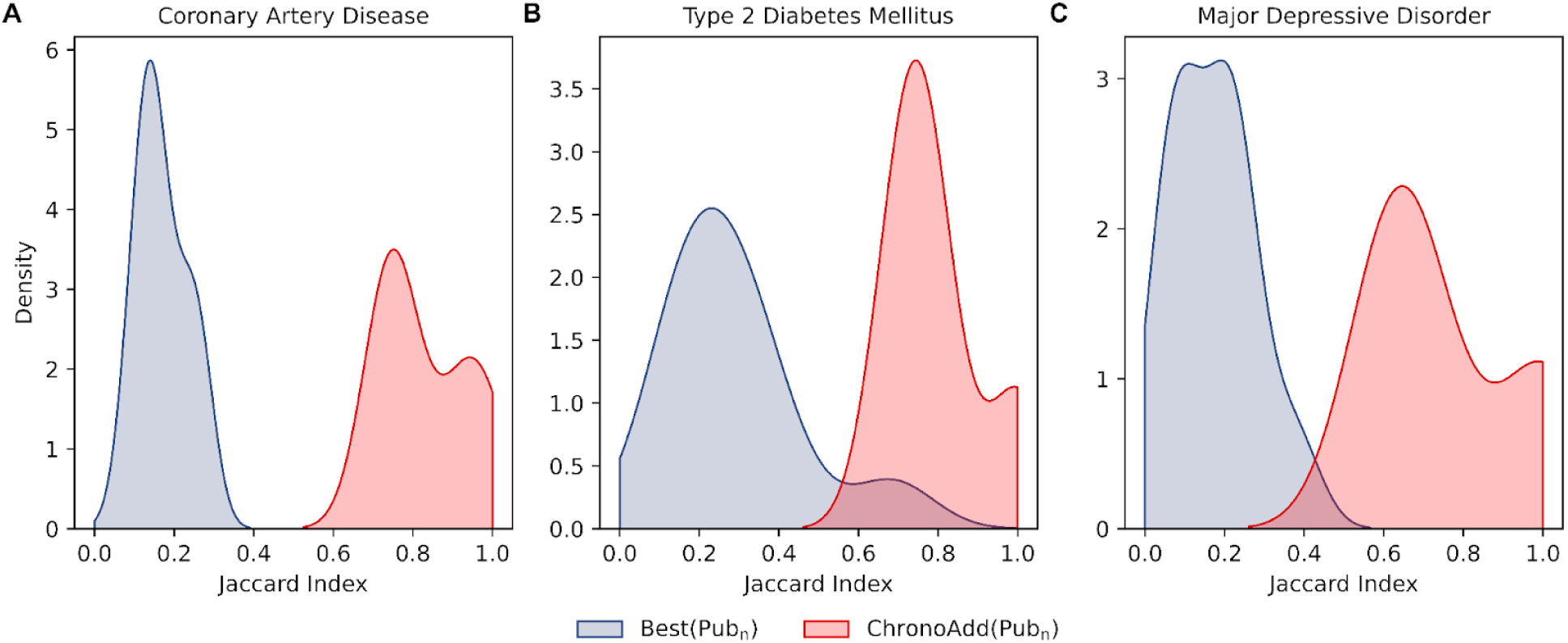
Jaccard index distributions for high genetic risk determined by trait-specific Best(Pub_n_) vs. ChronoAdd(Pub_n_) from five most recent publications. Jaccard indices of similarity calculated for pairs of classification of top 10% risk determined by trait-specific polygenic scores from the last five respective publications. Best(Pub_n_) indicates the single best trait-specific PRS from each of the last 5 publications, ordered by date of publication. ChronoAdd(Pub_n_) indicates polygenic scores generated using the PRSmix framework, wherein all scores from each Pub_n_ were combined with all the scores from all other Pub_n_ published before it using PRSmix.

**Figure 6:**
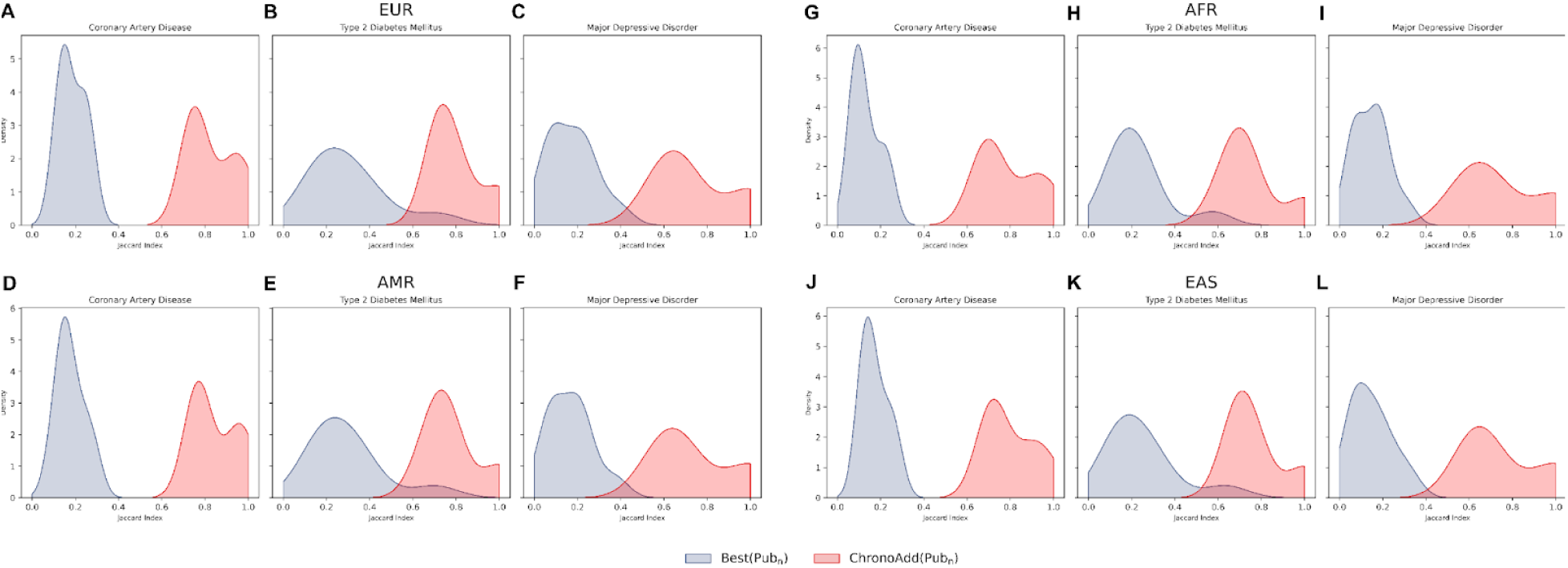
Jaccard index distributions for high genetic risk determined by trait-specific Best(Pub_n_) vs. ChronoAdd(Pub_n_) from five most recent publications across genetically predicted ancestry groups. Jaccard indices of similarity calculated for pairs of classification of top 10% of risk determined by trait-specific polygenic scores from the last five respective publications. Best(Pub_n_) indicates the single best trait-specific PRS from each of the last five publications, ordered by date of publication. ChronoAdd(Pub_n_) indicates polygenic scores generated using the PRSmix framework wherein all scores from each Pub_n_ were combined with all the scores from all other Pub_n_ published before it using PRSmix. Genetically inferred ancestry based on k-nearest neighbor approach as European (EUR), African (AFR), admixed American (AMR), and East Asian (EAS).

As a result of the greater congruence in high-risk classification, using ChronoAdd(Pub_n_) led to more consistent estimation of genetic risk percentile for individual participants. Individuals classified in top 10% risk by the ChronoAdd(Pub_Last_) for CAD have median percentiles across Best(Pub_n_) of 86 [79-91.5] vs. 93 [87-97] for ChronoAdd(Pub_n_).(Figure 7A) Similarly, individuals classified in the top 10% risk by the ChronoAdd(Pub_Last_) for T2DM have median percentiles across Best(Pub_n_) of 87 [81-93] vs. 95 [91-98] for ChronoAdd(Pub_n_).(Figure 7B) Additionally, individuals classified in the top 10% risk by the ChronoAdd(Pub_Last_) for MDD have median [IQR] percentiles across Best(Pub_n_) of 86 [79-92] vs. 95 [92-98] for ChronoAdd(Pub_n_). (Figure 7C) For any given Best(Pub_n_), high-risk individuals fall to a lower, wider range of percentiles across all Best(Pub_n_) when compared with their paired ChronoAdd(Pub_n_) scores. Among individuals classified in the top 10% of risk by ChronoAdd(Pub_Last_) for each trait, the median [IQR] percentile according to the Best(Pub_n_) that had the most inconsistent high-risk classification relative to ChronoAdd(Pub_Last_) out of the last five publications, was 78 [55-92] vs. 95 [92-98] in its paired ChronoAdd(Pub_n_) score for CAD. Similarly, it was 87 [71-95] vs.96 [93-98] for T2DM, and 59 [33-81] vs. 96 [93-98] for MDD.(Figure 7). These findings of improved individual-level classification for high genetic risk of CAD, T2DM, and MDD generalize across ancestral subgroups.(Figure 8).

**Figure 7:**
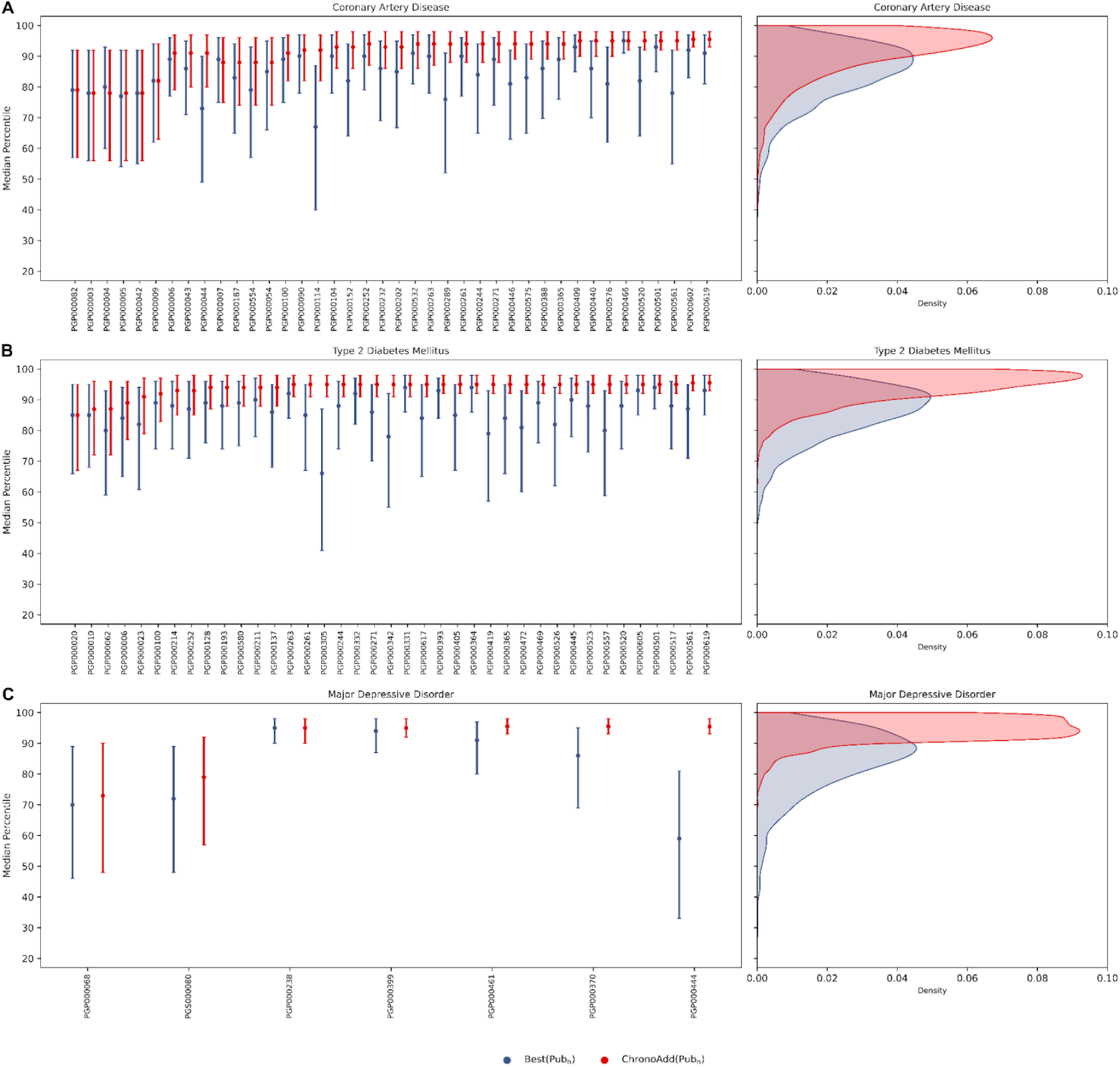
Median percentile and score-specific percentile distributions of Best(Pub_n_) vs. ChronoAdd(Pub_n_) across three traits for individuals identified as having high genetic risk by ChronoAdd(Pub_Last_) Distributions of the median percentile per polygenic score type for individuals classified in the top 10% risk by the ChronoAdd(Pub_Last_) for each disease of interest. Best(Pub_n_) indicates the single best trait-specific PRS from each publication Pub_n_, ordered by date of publication. ChronoAdd(Pub_n_) indicates polygenic scores generated using the PRSmix framework, wherein all scores from each Pub_n_ were combined with all the scores from all other Pub_n_ published before it using PRSmix. CAD: coronary artery disease; T2DM: type 2 diabetes; MDD: major depressive disorder. A) Individuals classified in top 10% risk by the ChronoAdd(Pub_Last_) for CAD have median percentiles across Best(Pub_n_) of 86 [79-91.5] vs. 93 [87-97] for ChronoAdd(Pub_n_). B) Similarly, individuals classified in the top 10% risk by the ChronoAdd(Pub_Last_) for T2DM have median percentiles across Best(Pub_n_) of 87 [81-93] vs. 95 [91-98] for ChronoAdd(Pub_n_). C) Additionally, individuals classified in the top 10% risk by the ChronoAdd(Pub_Last_) for MDD have median percentiles across Best(Pub_n_) of 86 [79-92] vs. 95 [92-98] for ChronoAdd(Pub_n_)_n_.

**Figure 8:**
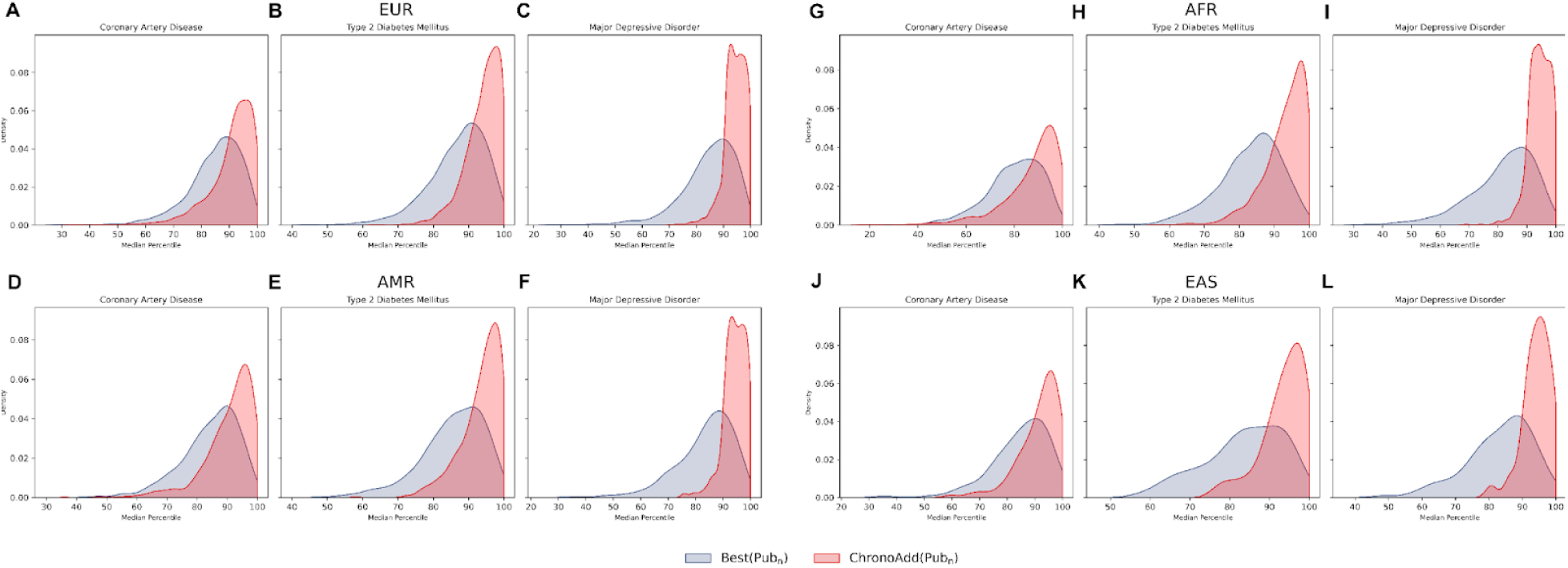
Median percentile distributions of Best(Pub_n_) vs. ChronoAdd(Pub_n_) across three traits for individuals identified as having high genetic risk by ChronoAdd(Pub_Last_), across genetically predicted ancestry groups. Distributions of the median percentile per polygenic score type for individuals classified in top 10% risk by the ChronoAdd(Pub_Last_) for each disease of interest. Best(Pub_n_) indicates the single best trait-specific PRS from each publication Pub_n_, ordered by date of publication. ChronoAdd(Pub_n_) indicates polygenic scores generated using the PRSmix framework, wherein all scores from each Pub_n_ were combined with all the scores from all other Pub_n_ published before it using PRSmix. Genetically inferred ancestry based on k-nearest neighbor approach as European (EUR), African (AFR), admixed American (AMR), and East Asian (EAS).

### Benchmarking associations with outcomes using ChronoAdd(Pub_n_) polygenic scores

Chronologically adding all scores from each publication Pub_n_ in order of publication into successive ChronoAdd(Pub_n_) scores led to progressively stronger associations with the outcome. The ORs were significantly stronger for the top 10% of individuals using ChronoAdd(Pub_n_) vs. Best(Pub_n_) respectively, with median [IQR] ORs for CAD (2.07 [1.77-2.25] vs. 1.68 [1.48-1.87]), T2DM (2.49 [2.39-2.53] vs. 1.86 [1.60-1.96]), and MDD (1.47 [1.37-1.49] vs. 1.30 [1.17-1.38]). This is particularly evident when restricting to the five most recent publications, with median [IQR] ORS for CAD (2.59 [2.58-2.59] vs. 1.90 [1.51-1.99]), T2DM (2.53 [2.53-2.54] vs. 2.30 [1.93-2.55]), and MDD (1.49 [1.47-1.49] vs. 1.32 [1.30-1.44]).(Figure 9A-C) We observed similar trends when comparing association strengths of Best(Pub_n_) vs. ChronoAdd(Pub_n_) in ancestral subgroups.(Supplemental Tables 4-6) Statistics for variance reflected this trend as well. The Nagelkerke’s pseudo-*R*² (median [IQR]) associated with top 10% risk classification was higher for ChronoAdd(Pub_n_) vs. Best(Pub_n_) in CAD (0.0089 [0.0053-0.0114] vs. 0.0044 [0.0024-0.0062]), T2DM (0.0176 [0.0158-0.0181] vs. 0.0076 [0.0043-0.0091]), and MDD (0.0033 [0.0022-0.0036] vs. 0.0014 [0.0005-0.0023]). This issue persists when restricting to the last five publications, with median [IQR] *R*² of (0.0162 [0.0161-0.0163] vs. 0.0070 [0.0025-0.0077]) in CAD, (0.0182 [0.0182-0.0184] vs. 0.0147 [0.0085-0.0186]) in T2DM, and (0.0036 [0.0033-0.0036] vs. 0.0017 [0.0014-0.0029]) in MDD.(Figure 9D-F) Similarly, the trends of increasing strength in ChronoAdd(Pub_n_) were additionally observed when looking at discrimination, where the C-statistics associated with the top 10% risk classification using ChronoAdd(Pub_n_) vs. Best(Pub_n_) when restricting to the last five publications ranged from 0.545-0.546 vs. 0.515-0.536 for CAD, 0.545-0.545 vs. 0.520-0.546 for T2DM, and 0.521-0.522 vs. 0.500-0.521 for MDD. (Figure 9G-I)

**Figure 9:**
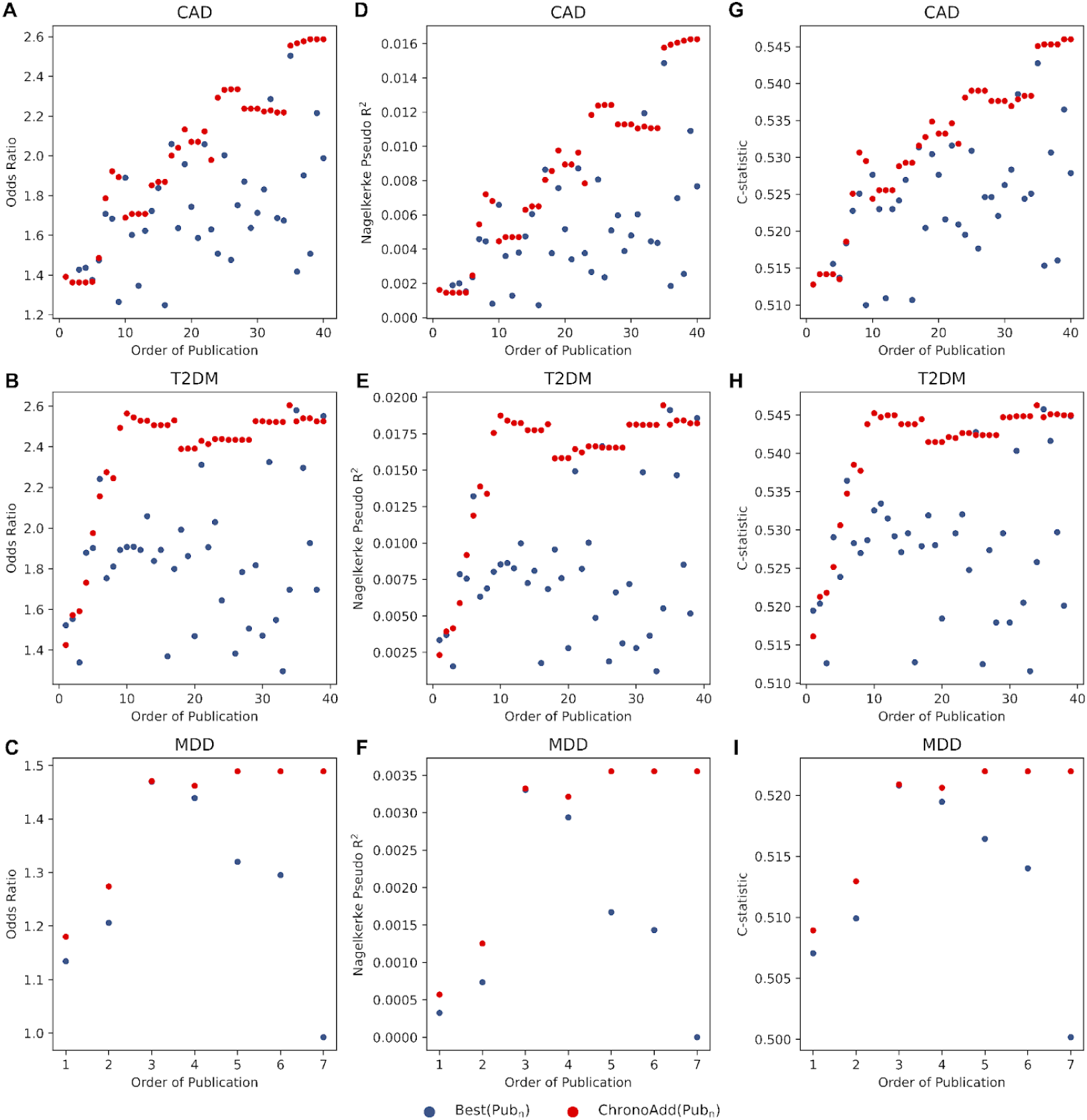
Chronologic trends in association strength, variance, and discrimination for Best(Pub_n_) vs. ChronoAdd(Pub_n_) across three traits. All publications for each trait were ordered chronologically by publication date. Best(Pub_n_) indicates the single best trait-specific PRS from each publication Pub_n_, ordered by date of publication. ChronoAdd(Pub_n_) indicates polygenic scores generated using the PRSmix framework, wherein all scores from each Pub_n_ were combined with all the scores from all other Pub_n_ published before it using PRSmix. High genetic risk is defined as the top 10% of each respective polygenic score. CAD: coronary artery disease; T2DM: type 2 diabetes; MDD: major depressive disorder. A-C) Odds ratios are based on logistic regression models predicting disease of interest, with variables of high genetic risk, age, sex, and first ten principal components of genetic ancestry. D-F) Nagelkerke’s pseudo-*R²* metric, as the difference of the full model inclusive of the polygenic score plus age, sex, and the first ten principal components of ancestry minus *R²* for the covariates alone. G-I) C-statistics are based on logistic regression models predicting disease of interest, with high genetic risk as the only variable. The proportion of phenotypic variance is explained by the high polygenic risk classification by each polygenic score for each respective disease.

## Discussion

In a comprehensive assessment of published PRS for three complex diseases, high PRS classification of individual participants was highly inconsistent across individual PRS. These findings are consistent with tail independence in extreme value theory seen with other multivariable distributions seen in clinical risk predictors. In the case of polygenic scores, because the strengths of association of individual PRS were significant at the population level, each score likely captures complementary predictive information as a by-product of its training process. This can be a function of the size of its GWAS, the composition of the training dataset, the exact phenotype definition, and assumptions of the polygenic score method, among other factors. We observed instability in risk estimates even between scores generated from the same GWAS or trained in the same cohort study. This instability builds on the inherent large variances in individual PRS estimates due to propagation of error as a function of GWAS sample size, number of causal SNPs and SNP-heritability – collectively resulting in instability of prediction across scores.^27^

Discrepancy in classification of high genetic risk is problematic in current clinical implementation efforts based on single PRS. As GWAS continue to grow and PRS methods continue to improve, newer PRS will inevitably be published. While we observe a high degree of correlation of scores across population-level metrics, we show that each individual published score has poor agreement of high-risk (as defined as being in the top 10%) status when compared with other individual scores. As PRS continue to demonstrate iterative improvement with newer publications, adoption of a new single PRS by a healthcare system followed by subsequent recalculation of risk with newer scores will lead to continual and significant reclassification of high-risk among patients, contributing to confusion and understandable lack of confidence in predicted risk. We observed significant variability in high-risk classification even among more recently published scores that used the same source genome-wide association studies for variant weights and reported similarly strong associations with phenotype in their publications. In addition to reporting population-based association, prediction, and calibration metrics, benchmarking high-risk classification by individual scores in hold-out datasets to understand these implications is critical prior to implementation.Use of integrative scoring to sequentially incorporate new PRS as they are published is a framework that can be deployed by biobanks to provide a more robust and stable classification of high-risk as well as improved overall PRS population-based metrics. This method excludes highly correlated scores and utilizes complementary information of available PRS to help predict risk more accurately in the target population. We showed that very significant gains in predictive performance are possible with additions of new, particularly strong PRS and that this leads to more substantial and likely more appropriate high-risk classification changes with this approach. For example, we observed several rises followed by plateaus in performance metrics which generally coincided with the incorporation of newer, larger, and more diverse GWAS datasets in PRS. Similarly, although smaller and statistically insignificant drops in performance metrics were also observed with additions of some scores related to differential prediction in our multi-ancestry testing cohort, high-risk classification more continually improved with newer ChronoAdd(Pub_n_) scores. Future iterations of the integrative scoring approach may also be applied to specific ancestry subpopulations.

When a new score is published, it can be incorporated into the integrative score model yielding an improved PRS with less marked variability in updating the classification of high-risk for individuals. Biobanks seeking to implement this approach would just need to identify new scores published for a target phenotype since the last PRS report was generated. Consideration of individual score performance metrics is not necessary as the elastic net approach will empirically reassess and weight component scores depending on new and orthogonal predictive information provided. There is nominal additional computational effort needed in calculating the input scores needed to calculate ChronoAdd scores. Moreover, newer iterations of ChronoAdd(Pub_n_) would be able to utilize the same clinical validation protocols prior to translation into patient care. Overall, the improved accuracy and consistency of high-risk classification between ChronoAdd(Pub_n_) scores and its streamlined implementation steps make ChronoAdd an ideal framework for incorporation into clinical workflows for PRS reporting and updating.

This study has several limitations. To mirror the development of the majority of published PRS, and given the recency of and middle-aged baseline of AOU, we performed association analyses with prevalent disease and still found significant instability in prediction. Future efforts on instability of incident disease prediction with follow-up beginning early in life will be additionally informative. We also used hospitalization and procedural codes to classify disease instead of clinically adjudicated outcomes. In this manuscript, we focus on common complex diseases for well-powered analyses. While some reports use OR for high-risk classification, we show wide variability in population-level metrics and percentile classification leading to wide variability in individual-level OR estimation by Best(Pub_n_). Thus, the proposed integrative scoring approach provides stability for both percentile and OR-based reporting.

In conclusion, in a comprehensive assessment of published PRS for three complex diseases, high PRS classification was highly inconsistent. Use of integrative scoring via PRSmix to sequentially incorporate new PRS enables standardization toward more stable and reliable classification of high risk.

## Methods

### Study population

The *All of Us* (AOU) Research Program is a cohort study focused on recruiting individuals traditionally underrepresented in biomedical research. Since 2018, AOU has enrolled people aged 18 and older from over 730 sites across the United States. The program has consented more than 800,000 participants, of whom over 560,000 have completed the basics of enrollment including collection of health questionnaires and biospecimens. For these participants, there is ongoing linkage to electronic health record (EHR) data, including ICD-9/ICD-10, SNOMED, and CPT codes. Genetic data comprises array samples from 315,000 participants and whole-genome sequencing (WGS) from 245,394 participants. This study used WGS data from the Controlled Tier Dataset version 7 release.

### Outcome ascertainment

CAD was defined based on self-report, occurrences of at least 2 diagnosis codes for myocardial infarction or a single procedure code for coronary revascularization. T2DM was defined based on diagnosis codes, laboratory results and medication prescriptions, as previously described by the eMERGE consortium.^28^ MDD was defined based on the presence of at least two diagnosis codes for major depressive disorder. All phenotype definitions are detailed in Supplemental Tables 9-11.

### Genotyping and quality control

Participants in AOU were genotyped using the Illumina Global Diversity Array at AOU genome centers. Central quality control measures included filtering for sex concordance, a cross-individual contamination rate below 3%, and a call rate above 98%. Further quality control performed by AOU included filtering for variants with population-specific allele frequency greater than 1% or a population-specific allele count greater than 100 in any AOU-computed ancestry subpopulations. Ancestry was inferred based on genetic similarity with projections of 20 principal components of genetic ancestry using 1000 Genomes as a reference panel. Inferred genetic ancestry in AOU was estimated in high-quality WGS samples that were restricted to bi-allelic sites, a minor allele frequency above 0.1%, a call rate above 99%, and a linkage disequilibrium-pruned threshold *R*² = 0.1.

### Polygenic score calculation

The variant effect sizes for 61 scores for CAD, 133 scores for T2DM and 18 scores for MDD were downloaded from Polygenic Score Catalog on May 28, 2024. Of these, we excluded 4 CAD scores and 4 T2DM scores that were already developed using PRSmix to prevent potential overfitting. This left 57 CAD scores, 129 T2DM scores, and 18 MDD scores for use in further analysis, none of which were previously trained in AOU or developed using AOU GWAS data. For score accession numbers see Supplemental Tables 1-3 and for source of variant annotations, see Supplemental Tables 12-14. All scores were harmonized to the AOU reference dataset using PRSmix. Scores with an OR/SD < 1 in the entire AOU cohort were considered anti-correlated and were thus inverted in all further analyses. All scoring in AOU was done using PLINK2 software. All polygenic scores were residualized for the first ten principal components of genetic ancestry and then standardized to have a mean of 0 and a standard deviation of 1 for each ancestry. Individuals were next binned into 100 groupings according to percentile of score, and individuals in the top 10% of the score distribution were deemed to have high genetic risk.

### Integration of polygenic scores using PRSmix

PRSmix is a framework that evaluates and leverages the data from a group of PRS for a target trait to generate a new score with improved prediction accuracy. PRSmix uses an elastic net model to produce a weighted linear combination of all the input PRS. The PRSmix framework was used first to harmonize all scores from the PGS Catalog to the AOU reference dataset via the *harmonize_snpeffect_toAL*T function. In addition, PRSmix was used to combine PRS via the *combine_PRS* function. PRSmix was run using all default parameters. This includes the use of age, sex, and the first 10 principal components as covariates, as well as an 80% vs. 20% split of the cohort into the training and testing cohorts, respectively.(Figure 1)

We set up a framework to compare predictive power and high-risk classification by individual vs. integrated polygenic scores. We ordered trait-specific polygenic score publications in the PGS catalog chronologically based on the source publication date, renaming them as Pub_n_, with n referring to the chronological order of publication. For each publication we tested each of its published score’s strength of association with the outcome of interest in the training cohort and labeled the strongest score as its corresponding Best(Pub_n_). This selection mechanism resulted in 40 Best(Pub_n_) out of 57 total CAD scores, 39 Best(Pub_n_) out of 129 total T2DM scores, and 7 Best(Pub_n_) out of 18 total MDD scores. We then calculated the corresponding list of integrated scores using an temporal incorporation approach in the training cohort. We calculated a ChronoAdd(Pub_n_) score for each publication Pub_n_, wherein all trait-specific scores from the given Pub_n_ as well as all scores from all publications prior to it were combined using PRSmix.

This process resulted in 40 ChronoAdd(Pub_n_) CAD scores, 39 ChronoAdd(Pub_n_) T2DM scores, and 7 ChronoAdd(Pub_n_) MDD scores. The Best(Pub_n_) and ChronoAdd(Pub_n_) scores were paired and directly compared for subsequent analyses with respect to order of publication. For each trait, the last version of the ChronoAdd(Pub_n_) score was labeled as the ChronoAdd(Pub_Last_) score. The top 10% of the ChronoAdd(Pub_Last_) distribution was used to define individuals with true high genetic risk for benchmarking studies.

### Statistical analysis

The association of polygenic scores with outcome of interest were assessed using logistic regression with covariates of enrollment age, sex, and the first 10 principal components of genetic ancestry. The discrimination of each of these polygenic scores was assessed using Harrell’s C-statistic. The proportion of phenotypic variance explained by the polygenic score on the observed scale was determined using Nagelkerke’s pseudo-*R²* metric via the rcompanion R package. This calculation involved finding the *R²* for the complete model, which included the variable of interest and baseline model covariates, and then subtracting the *R²* for the baseline covariates alone. Congruence of risk estimates was assessed with the Jaccard similarity index, which measures the amount of overlap or similarity between two sample sets.^29^ Simulations of correlated distributions were performed in R. True risk was a randomly generated standard normal distribution for 10,000 data points, i.e. rnorm(10000). For each of the three expected correlation coefficients *r* of 0.2, 0.5, and 0.8, the 50 scores were simulated by permuting the true risk such that the correlation between the score and the true risk would be approximately *r*, i.e. *r* * true_risk + sqrt(1 – *r²*) * rnorm(10000) in R. All analyses were two-sided. Statistical analyses were performed using R version 4.3.1.

### Data availability

All data are made available from the *All of Us* Research Study to researchers from universities and other institutions with genuine research inquiries following institutional review board and *All of Us* approval. This research was approved by the Mass General Brigham institutional review board. The weights of the polygenic scores analyzed in this study are publicly available for download from the Polygenic Score Catalog.^23^ The mixing weights of the ChronoAdd(Pub_n_) are available in the Supplemental Tables 1-3.

## Acknowledgements

This work was supported by grants K08HL161448 (to A.C.F.), R01HL164629 (to A.C.F.), K08HL168238 (to A.P.P.), R01HL169015 (to A.P.P.), R01HL1427 (to P.N.), R01HL148565 (to P.N.), R01HL148050 (to P.N.) from the National Heart, Lung, and Blood Institute and grants T32HG010464 (to S.M.U.), R01HG012354 (to A.P.P.) and U01HG011719 (to A.P.P. and P.N.) from the National Human Genome Research Institute. This research has been conducted using the *All of Us* cohort study. We gratefully acknowledge *All of Us* participants for their contributions, without whom this research would not have been possible. We also thank the National Institutes of Health’s *All of Us* Research Program for making available the participant data examined in this study.

